# Risk Factors for Delirium in Elderly Patients After Lumbar Spinal Fusion

**DOI:** 10.1101/2022.01.20.22269610

**Authors:** Colin Gold, Emanuel Ray, David Christianson, Brian Park, Ioannis A Kournoutas, Taimur A Kahn, Eli A. Perez, Joel I. Berger, Katie Sander, Cassim A. Igram, Andrew Pugely, Catherine R. Olinger, Ryan Carnahan, Pei-fu Chen, Rashmi Mueller, Patrick Hitchon, Matthew A. Howard, Matthew Banks, Robert D. Sanders, Royce W. Woodroffe

## Abstract

**Background Context:** Postoperative delirium is a clinically significant acute disorder of consciousness especially prevalent in older adult patients, of which more than 100,000 per year undergo spinal fusion surgery. There are no proven preventative therapies, and delirium is associated with increased postoperative complications, functional decline, morbidity, and mortality.

**Purpose:** To identify perioperative risk factors for postoperative delirium (POD) after lumbar spinal fusion procedures in patients aged 65 or older.

**Study Design:** Retrospective Review

**Patient Sample:** 702 patients who underwent lumbar spinal fusion surgery from 11/13/2017 to 1/30/2021.

**Outcome Measures:** The primary outcome was the presence of postoperative delirium assessed by the Delirium Observation Screening Scale (DOSS) and Confusion Assessment Method for the ICU (CAM-ICU).

**Methods:** Demographic, surgical, and perioperative data were obtained from the electronic medical records. The primary outcome was presence of postoperative delirium. Univariate and multivariate analyses were performed. A binomial logistic regression model was designed using a custom written MATLAB script.

**Results:** Of the 702 patients included in the study, 173 (24.6%) developed POD. Our analysis revealed that older age (*p* < 0.001), lower preoperative hemoglobin (*p* < 0.001), and higher ASA grade (*p* < 0.001), were significant preoperative risk factors for developing POD. The only significant intraoperative risk factor was a higher number of spinal levels that were instrumented (*p* < 0.001). Higher pain scores on postoperative day 1 (*p* < 0.001), and lower postoperative hemoglobin (*p* < 0.001) were associated with increased POD; as were ICU admission (*p* < 0.001) and increased length of ICU stay (*p* < 0.001). Patients who developed POD had a longer hospital stay (*p* < 0.001) with lower rates of discharge to home as opposed to an inpatient facility (*p* < 0.001).

**Conclusions:** Risk factors for older adults undergoing lumbar spinal fusion surgery include advanced age, lower preoperative and postoperative hemoglobin, higher ASA grade, greater extent of surgery, and higher postoperative pain scores. Patients with delirium had a higher incidence of postoperative ICU admission, increased length of stay, decreased likelihood of discharge to home and increased mortality, all consistent with prior studies. Further studies will determine whether adequate management of anemia and pain lead to a reduction in the incidence of postoperative delirium in these patients.

## INTRODUCTION

Delirium is an abrupt disturbance of cognition and consciousness and is characterized by a decreased ability to sustain or shift attention, impairment of memory and executive function, and fluctuation in arousal levels.[1] The incidence of delirium after surgery has been reported to be approximately 20% in patients having spine surgery,[2-4] and as high as 50-80% in critically ill patients. ^2-4^ It is especially prevalent in the elderly population with almost a 15-fold increase in patients over the age of 70 years.[5] Postoperative delirium, with its associated high rate of other complications, functional decline, and mortality, inflicts an immense burden on patients, the healthcare industry and society.[6-8] In 2008, the annual financial burden due to delirium in elderly hospitalized patients was estimated to be up to $152 billion in the USA.[9] Prolonged length of hospitalization in older patients with postoperative delirium contributed in a large measure to this expenditure.^9^ In the intervening 12 years, this problem has almost certainly been further exacerbated by a growing elderly population and poses a major challenge. Currently, there is poor understanding of the underlying mechanisms of delirium.[10, 11] However, previous research suggests that some of the factors that delirium is associated with include preexisting poor functional status[12], neuroinflammation,[13] a breakdown of the blood-brain barrier,[14] and intraoperative blood loss.[15] While there have been studies examining risk factors for postoperative delirium, to our knowledge, none have specifically determined perioperative risk factors after lumbar spinal fusion. The primary objective of this study was to identify the incidence of, the risk factors for, and the consequences of, postoperative delirium in geriatric patients undergoing lumbar spinal fusion.

## PATIENTS AND METHODS

### Data source and patient cohort

After obtaining Institutional Review Board approval (IRB No. 202007546), we queried our institution’s electronic medical records for patients who were ≥65 years of age and underwent elective posterior thoracolumbar or lumbosacral fusion between November 2017 and January 2021. The procedures were performed both by neurosurgeons and orthopedic surgeons. A total of 708 patients were identified. Only patients who had delirium assessment screening performed while in the hospital were included (n= 702); those without such assessments were excluded from the analysis (n=6).

Demographic information including age, gender, body mass index (BMI) and data about the presence of baseline medical comorbidities including anemia, diabetes, hypertension, as well as the preoperative functional status and degree of preexisting illness as assessed by the American Society of Anesthesiologists (ASA) physical status was collected. Additional data including the duration of the procedure from incision to skin closure, length of time under anesthesia, estimated blood loss (EBL), number of units of blood transfused, preoperative and postoperative hemoglobin levels, intraoperative administration of tranexamic acid (TxA) and dexmedetomidine, and the use of intraoperative neurophysiological monitoring using motor evoked potentials were also collected. The extent of surgical intervention was quantified by the number of vertebral levels instrumented and the number of intervertebral discs that were removed and replaced by interbody grafts. The length of time under anesthesia was based on the times charted by the operating room nurse in the medical record for “Anesthesia Start” and “Anesthesia Stop.” These usually correlated to the time prior to the induction of general anesthesia but after the preinduction safety checklist had been reviewed, and the exit of the patient from the operating room, respectively. Postoperative data gathered were the need for intensive care unit (ICU) admission, length of ICU stay, pain scores using the Visual Analogue Scale (VAS), and the dosage of gabapentin, and opioids (morphine equivalent dosage) administered.

The primary endpoint of delirium was assessed by the Delirium Observation Screening Scale (DOSS) and the Confusion Assessment Method for the ICU (CAM-ICU).[16, 17] At our institution these are routinely collected twice a day by the nursing staff on the ward. DOSS is used for non-intubated patients. Previous validation of assessments using DOSS by nurses at our institution demonstrated that scores ≥3 were 90% sensitive and 91% specific for delirium.[18] CAM-ICU is used for intubated patients and was 95-96% sensitive and 93% specific when administered by nurses in the original validation study.[17]

Data for the first seven days after surgery was collected and analyzed. Delirium was represented by a categorical “yes” or “no” for the presence of delirium; the number of days that delirium was present was estimated as an integer variable. Patients with incomplete screening data were presumed not to be delirious on days for which data was missing (n=24). Secondary outcomes included the overall length of hospitalization and the disposition at discharge -to their home, an acute rehabilitation facility, a skilled nursing facility, or a long-term acute care hospital.

### Statistical analysis

Descriptive statistics were used to represent demographic and intraoperative data. Patients’ VAS pain scores were averaged by their postoperative day. Categorical variables were summarized using frequencies, and proportions were compared using Pearson’s Chi-square test when applicable or a Fisher’s Exact Test when the expected cell sizes were small. Continuous variables were summarized by means and standard deviation, and were compared using unpaired, two-tailed Welch’s t-tests or analysis of variance (ANOVA) when comparing more than 2 groups. The results of the tests listed above were used to guide variable selection for pooled logistic regression models in which the presence of delirium was the dependent variable. An initial model (Model 1) included data up to and including the day a patient became delirious (in the scenario where this event occurred). Although this model is helpful for including all available and potentially explanatory data, certain variables could be confounded by the presence of delirium at that final data point (i.e. the day of delirium), wherein delirium itself could be a causal factor for a change in those variables. Therefore, we also ran an additional model (Model 2) which excluded data from patients on the day they became delirious. Variables with a high range (values >100) were log_2_-transformed prior to input into the models, to aid with interpreting the output. In this scenario, odds ratios (OR) relate to a doubling of a particular value rather than a single unit of change. Statistical analysis was performed in GraphPad Prism (version 9.0, GraphPad Software) and IBM SPSS Statistics (version 25.0, IBM Corp.). Logistic regression models were run in MATLAB (MathWorks Inc.) using the *fitglm* function. Statistical significance was established with an alpha 0.001 after Bonferroni correction (0.05/44).

## RESULTS

All 702 patients who were 65 years of age or older had DOSS or CAM-ICU data available. From this sample, 173 patients screened positive for delirium on at least one day in the first week following surgery (24.6%). In 118 of the 173 patients, 68.2%, delirium was noted on more than one day (Figure 1). In 42.8% of patients, the onset of delirium was observed on postoperative day 1 (Figure 2). The average onset of delirium was 2.2 days after surgery and the average duration of delirium was 2.8 days.

**Figure 1.**
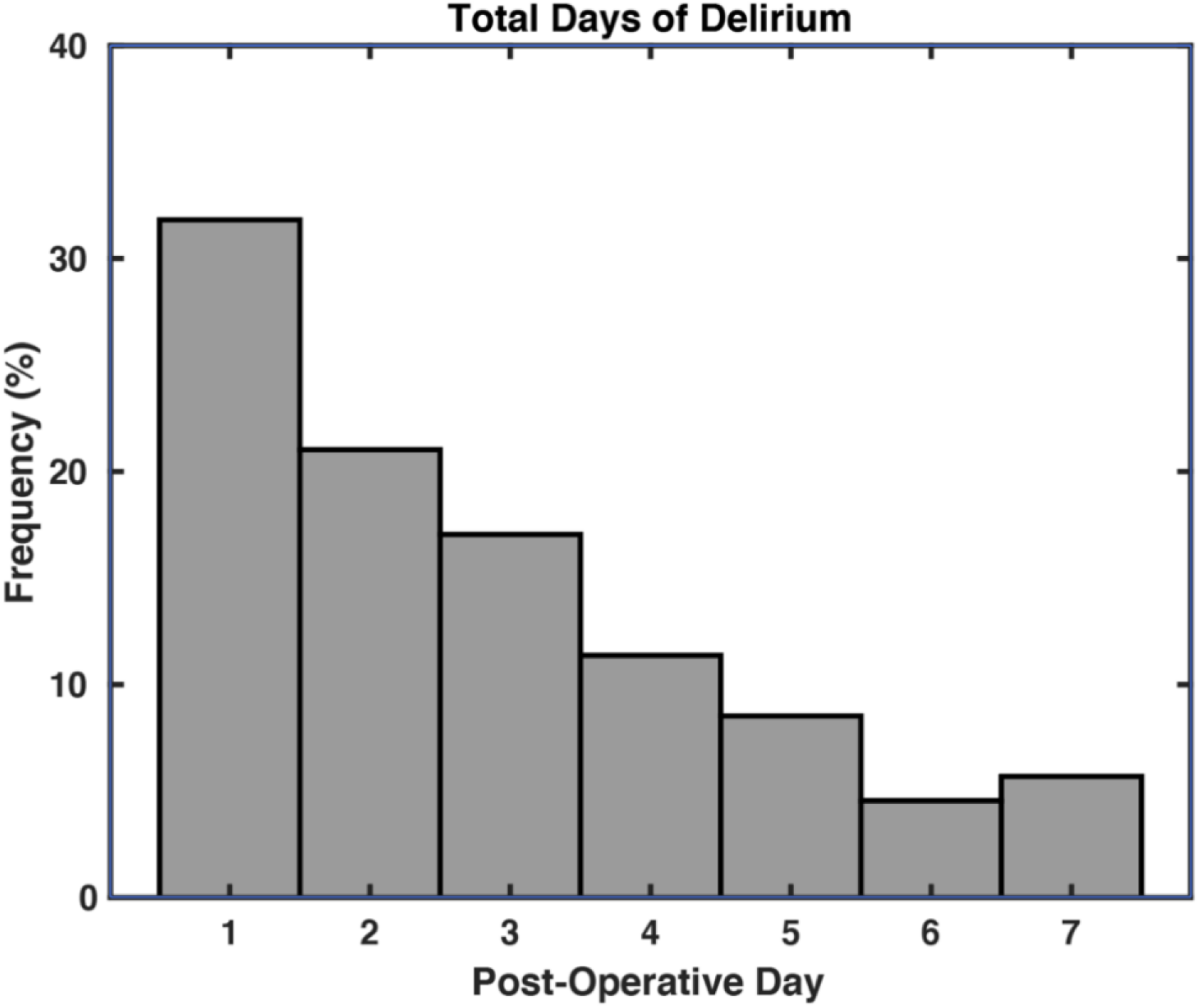
Frequency histogram of total days of delirium for each patient who developed delirium.

**Figure 2.**
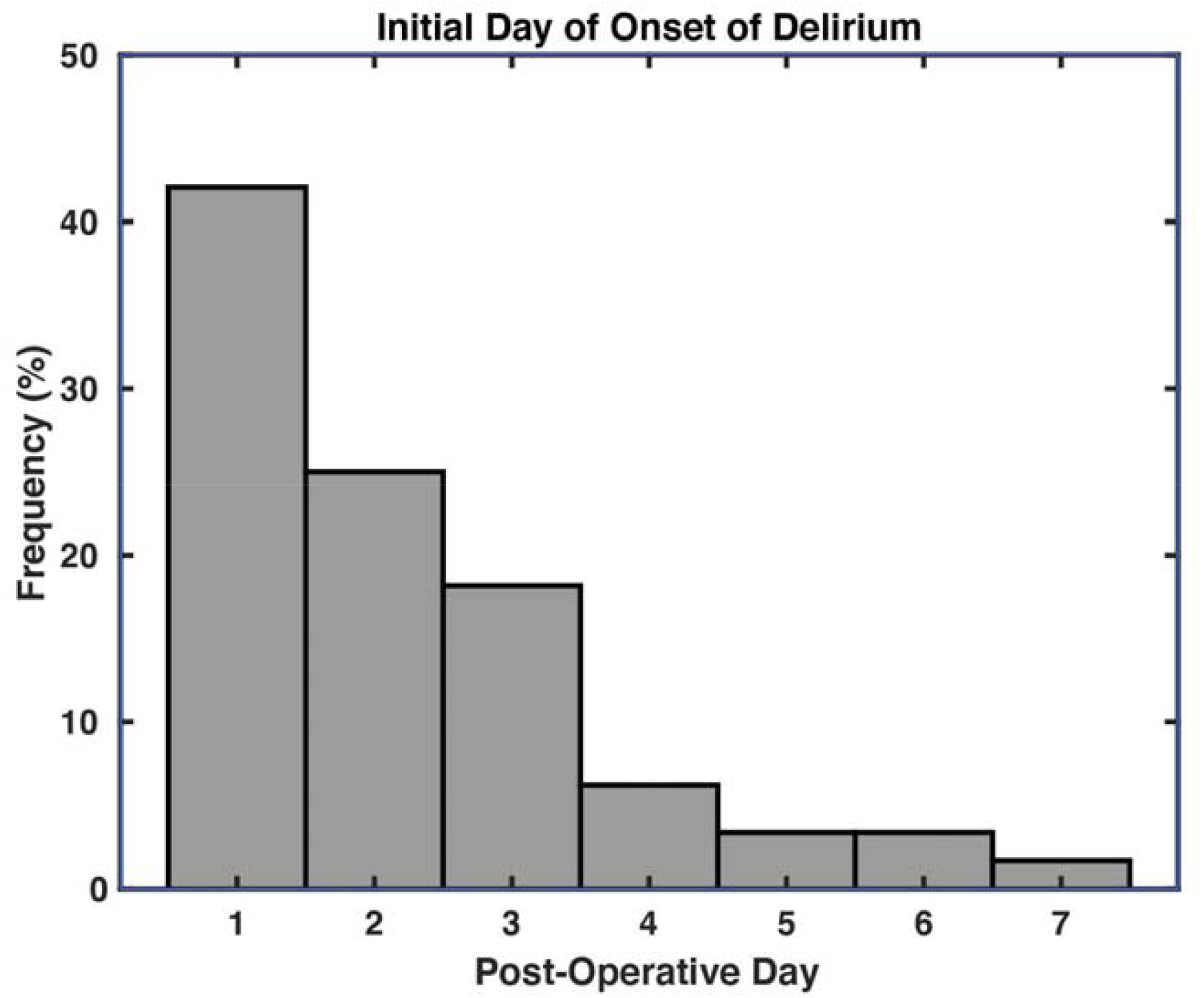
Frequency histogram of initial day of delirium onset for each patient who developed delirium.

Advanced age was associated with delirium (Table 1); delirious patients had a mean age of 73.8 years, and the mean age for non-delirious ones was 71.7 (*p* < 0.001). Gender had no significant association to delirium (*p* = 0.089). The ASA physical status was associated with delirium based on one-way ANOVA (*p* < 0.001). Only two patients had an ASA score of 1 and neither developed delirium. Of the 280 patients, 16.1% with an ASA score of 2 developed delirium, 30.1% of the 399 patients who were ASA score 3 developed delirium, and 40.0% of the 20 patients who were ASA grade 4 developed delirium. Diabetes was not significantly associated with delirium after Bonferroni correction (*p* = 0.005). The incidence of delirium was 33.1% in diabetic patients and 22.1% in non-diabetic patients (*p* = 0.005). Preexisting dementia or mild cognitive impairment was not significantly associated with delirium; however, only 3 patients had such a diagnosis and 2 of them developed postoperative delirium (*p* = 0.091). The number of levels involved in posterior instrumentation significantly correlated with delirium. A mean number of 4.9 levels were instrumented in the delirium group and 4.0 (*p* < 0.001) in the non-delirious group. The number of interbody grafts placed was not significantly associated with delirium (*p* = 0.118).

**Table 1.** Patient demographic information stratified by Delirium.

|  | Overall<br>(n = 702) | Postoperative delirium<br>(n=173) | No postoperative<br>delirium (n = 529 ) | p-value* |
| --- | --- | --- | --- | --- |
| **Age, mean (SD)** | 72.2 (5.3) | 73.7 (5.9) | 71.7 (5.0) | **<0.001** |
| **Gender** |  |  |  |  |
| Male | 364 | 80 (22.0 %) | 284 (78.0%) | 0.089 |
| Female | 338 | 93 (27.5 %) | 245 (72.5 %) |  |
| **BMI, mean (SD)** | 31.0 (11.4) | 30.1 (6.8) | 31.3 (12.5) | 0.359 |
| **Diabetes** | 160 | 53 (33.1%) | 107 (66.9%) | 0.005 |
| **Hypertension** | 386 | 103 (26.7 %) | 283 (73.3 %) | 0.166 |
| **ASA Status** |  |  |  | **<0.001\*** |
| Grade 1 | 2 (0.3%) | 0<sup>a</sup> | 2 (0.4%)<sup>a</sup> |  |
| Grade 2 | 280 (39.9%) | 45 (26.0%)<sup>a</sup> | 235 (44.5%)<sup>b</sup> |  |
| Grade 3 | 399 (56.9%) | 120 (69.4%)<sup>a</sup> | 279 (52.8%)<sup>b</sup> |  |
| Grade 4 | 20 (2.9 %) | 8 (4.6%)<sup>a</sup> | 12 (2.3%)<sup>a</sup> |  |
| **Number of levels of**<br>**posterior instrumentation**<br>**(SD)** | 4.2 (2.7) | 4.9 (2.9) | 4.0 (2.6) | **<0.001** |
| **Number of interbody**<br>**grafts placed (SD)** | 0.7 (1.0) | 0.8 (1.1) | 0.7 (0.9) | 0.118 |
| **Dementia or Cognitive**<br>**Impairment** | 3 | 2 (66.6 %) | 1 (33.3 %) | 0.091 |

\*Superscripts denote significant difference between rows. Delirium rates in ASA grade 2 and grade 3 differ from each other significantly, but do not differ from grade 1 and grade 4.

The presence of delirium was not significantly associated with the anesthesia time as defined here, or the operative time (from skin incision to close; *p* = 0.019 and *p* = 0.076, respectively). Mean operative time for the delirium group was 4 h and the mean anesthesia time was 5 h 49 minutes. In the non-delirium group, the mean operative time was 3 h 44 min and the anesthesia time was 5 h 33 min. The estimated blood loss (EBL) was not found to be significantly related to delirium with a mean of 580.3 ml in the delirium group and 452.1 ml in the non-delirium group (*p* = 0.019). Total postoperative gabapentin dosage did not significantly correlate with delirium (5,223 mg total dosage in the delirium group versus 4,069 mg in the non-delirium group, *p* = 0.003; Table 2).

**Table 2.** Surgical information stratified by Delirium.

|  | Overall<br>(n = 702) | Postoperative<br>delirium (n=173) | No postoperative delirium<br>(n = 529) | p-value* |
| --- | --- | --- | --- | --- |
| **Operative time,**<br>**minutes, mean (SD)** | 228 (95) | 240 (105) | 224 (91) | 0.076 |
| **Anesthesia time,**<br>**minutes, mean (SD)** | 337 (77) | 349 (74) | 333 (78) | 0.019 |
| **Tranexamic acid** | 230 | 58 (25.2%) | 172 (74.6%) | 0.806 |
| **EBL, mL, mean (SD)** | 483.7 (586.6) | 580.3 (613.7) | 452.1 (574.6) | 0.019 |
| **Transfusion required** | 249 | 61 (24.5%) | 188 (75.5%) | 0.947 |
| **Pre-op Hemoglobin,**<br>**mean (SD)** | 13.2 (1.8) | 12.6 (1.9) | 13.4 (1.7) | **<0.001** |
| **Post-op Hemoglobin,**<br>**mean (SD)** | 10.6 (1.8) | 10.0 (1.6) | 10.8 (1.8) | **<0.001** |
| **Difference in Hg,**<br>**mean (SD)** | 2.6 (1.6) | 2.6 (1.8) | 2.6 (1.5) | 0.680 |
| **Intraoperative**<br>**dexmedetomidine** | 48 | 12 (25%) | 36 (75%) | 0.953 |
| **ICU admission** | 170 | 71 (41.7%) | 99 (58.2%) | **<0.001** |
| **Duration of ICU**<br>**admission, hours,**<br>**mean (SD)** | 58.9 (52.8) | 79.7 (73.4) | 44.0 (20.4) | **<0.001** |
| **Preoperative**<br>**gabapentin use** | 481 | 116 (24.1%) | 365 (75.6%) | 0.632 |
| **Postoperative**<br>**gabapentin use** | 522 | 129 (24.7%) | 393 (75.3%) | 0.943 |
| **Total postoperative**<br>**Gabapentin Dose (mg)** | 4354 (3864) | 5223 (3748) | 4069 (3862) | 0.003 |
| **Length of stay, days**<br>**mean (SD)** | 5.8 (5.6) | 9.2 (8.4) | 4.7 (3.7) | **<0.001** |
| **Disposition** |  |  |  | **<0.001\*** |
| Home | 399 (56.8%) | 51 (29.5%)<sup>a</sup> | 347 (65.6%)<sup>b</sup> |  |
| Acute Rehab | 103 (14.7%) | 35 (20.2%)<sup>a</sup> | 68 (12.8%)<sup>b</sup> |  |
| Skilled Nursing Facility | 190 (27.1%) | 78 (45.1%)<sup>a</sup> | 112(21.2%)<sup>b</sup> |  |
| Death/ Comfort Care | 3 (0.4%) | 2 (1.2%)<sup>a</sup> | 1 (0.2%)<sup>a</sup> |  |
| LTACH | 7 (1.0%) | 6 (3.5%)<sup>a</sup> | 1 (0.2%)<sup>b</sup> |  |
| **Postoperative surgical**<br>**site infection** | 30 | 18 (60%) | 12 (30%) | **<0.001** |
| **Reoperation required** | 100 | 20 (20%) | 80 (80%) | 0.371 |
| **Death within 90 days** | 14 | 10 (71.4%) | 4 (28.6%) | **<0.001** |

\*Superscripts denote columns that differ significantly from one another.

The difference between the preoperative and first postoperative hemoglobin levels was not associated with delirium (*p* = 0.680). However, both the preoperative (*p* < 0.001) and first postoperative (*p* < 0.001) hemoglobin levels were lower in delirious patients. The delirium group had a mean starting hemoglobin of 12.6, while the non-delirium group had a mean starting hemoglobin of 13.4. Both groups had an average decrease of 2.6 in their hemoglobin values. The average length of hospitalization was 9.2 days for the delirium group and 4.7 days for the non-delirium group (*p* < 0.001; Table 2). The rate of admission to the ICU and the duration of time spent in the ICU were also significantly higher in the delirium group (*p* < 0.001). Additionally, patients who developed POD were significantly less likely to be discharged home (29.5% vs. 65.6%), more likely to develop a surgical site infection (10.4% vs. 2.3%), and more likely to die within 90 days of surgery (5.7% vs. 0.7%).

The daily averages of pain scores reported by patients using the visual analog scale (VAS) were found to significantly correlate with the presence of delirium on postoperative day 1 (*p* < 0.001). In the delirium group, on day 1, the mean VAS scores were 5.01. In the non-delirium group, on day 1, the mean VAS score was 4.29. From days 2 to 7, average VAS scores did not significantly differ depending on the presence or absence of delirium (Figure 3). Additionally, the mean VAS score for the entire hospital stay was not different in the delirious versus non-delirious groups (*p* = 0.144; Table 3). The morphine equivalent dose (MED) of opioids administered was averaged for each postoperative day as well as cumulatively for the entire hospital stay. The mean MED was higher in the delirium group on each postoperative day except for day 7 (Figure 4). However, these differences were not statistically significant (Table 4).

**Figure 3.**
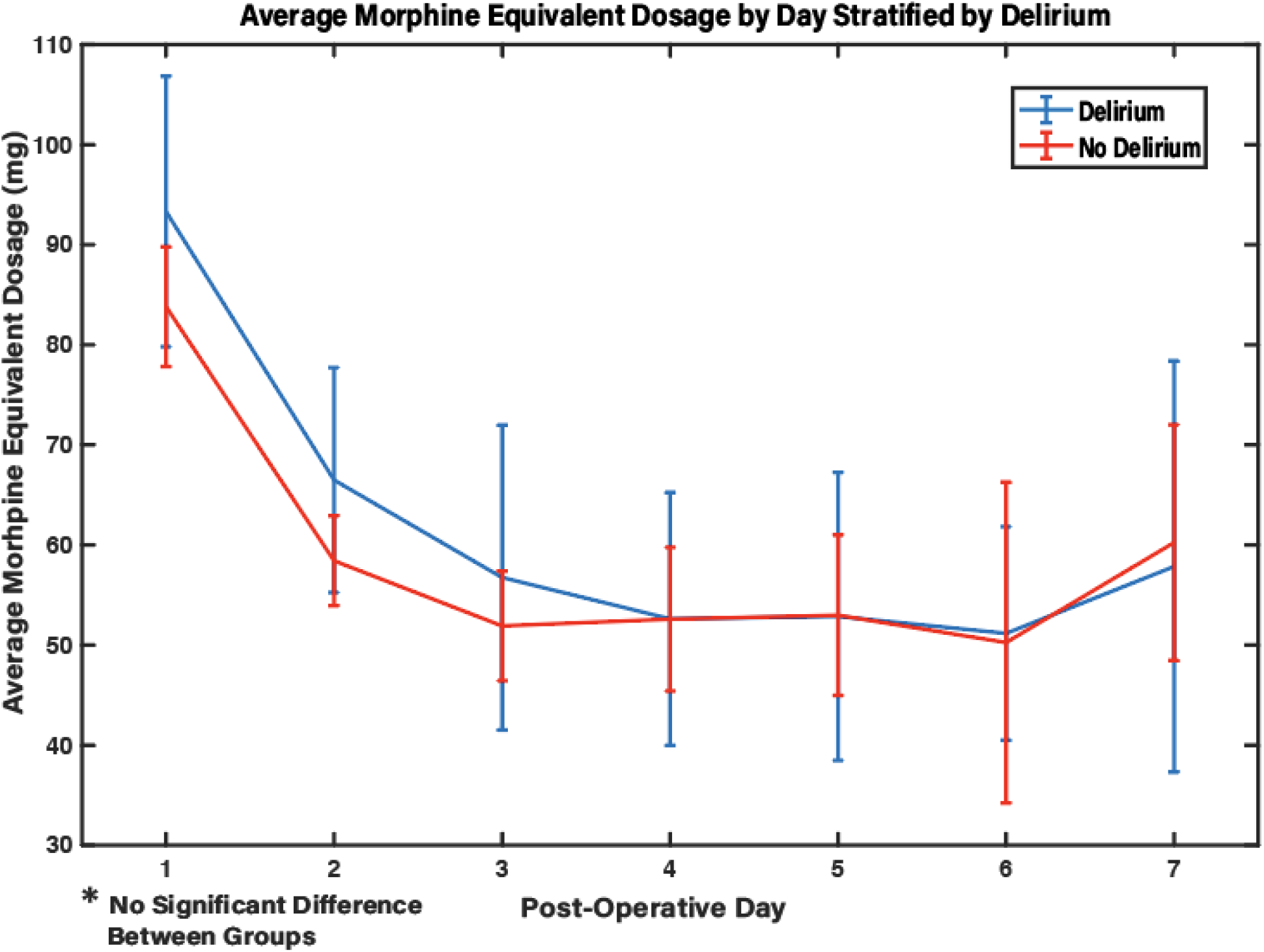
Line plot with 95% confidence intervals of average visual analog scale (VAS) pain score by postoperative day for delirious (blue) and non-delirious patients (red). The asterisk denotes significant difference between groups on postoperative day one.

**Table 3.** Mean pain scores compared between patients who developed delirium at any point during the postoperative period.

|  | Postoperative delirium | No postoperative delirium | p-value* |
| --- | --- | --- | --- |
| **VAS POD1, mean (SD)** | 5.01 (2.21) | 4.29 (1.75) | **<0.001** |
| **VAS POD2, mean (SD)** | 4.64 (2.20) | 4.17 (1.68) | 0.011 |
| **VAS POD3, mean (SD)** | 4.19 (2.30) | 4.09 (1.85) | 0.604 |
| **VAS POD4, mean (SD)** | 4.07 (2.40) | 4.10 (1.98) | 0.889 |
| **VAS POD5, mean (SD)** | 4.21 (2.25) | 3.88 (1.94) | 0.175 |
| **VAS POD6, mean (SD)** | 4.08 (2.34) | 4.16 (2.08) | 0.797 |
| **VAS POD7, mean (SD)** | 4.04 (2.61) | 4.50 (2.34) | 0.266 |
| **VAS during admission, mean (SD)** | 4.29 (1.83) | 4.08 (1.54) | 0.144 |

**Figure 4.**
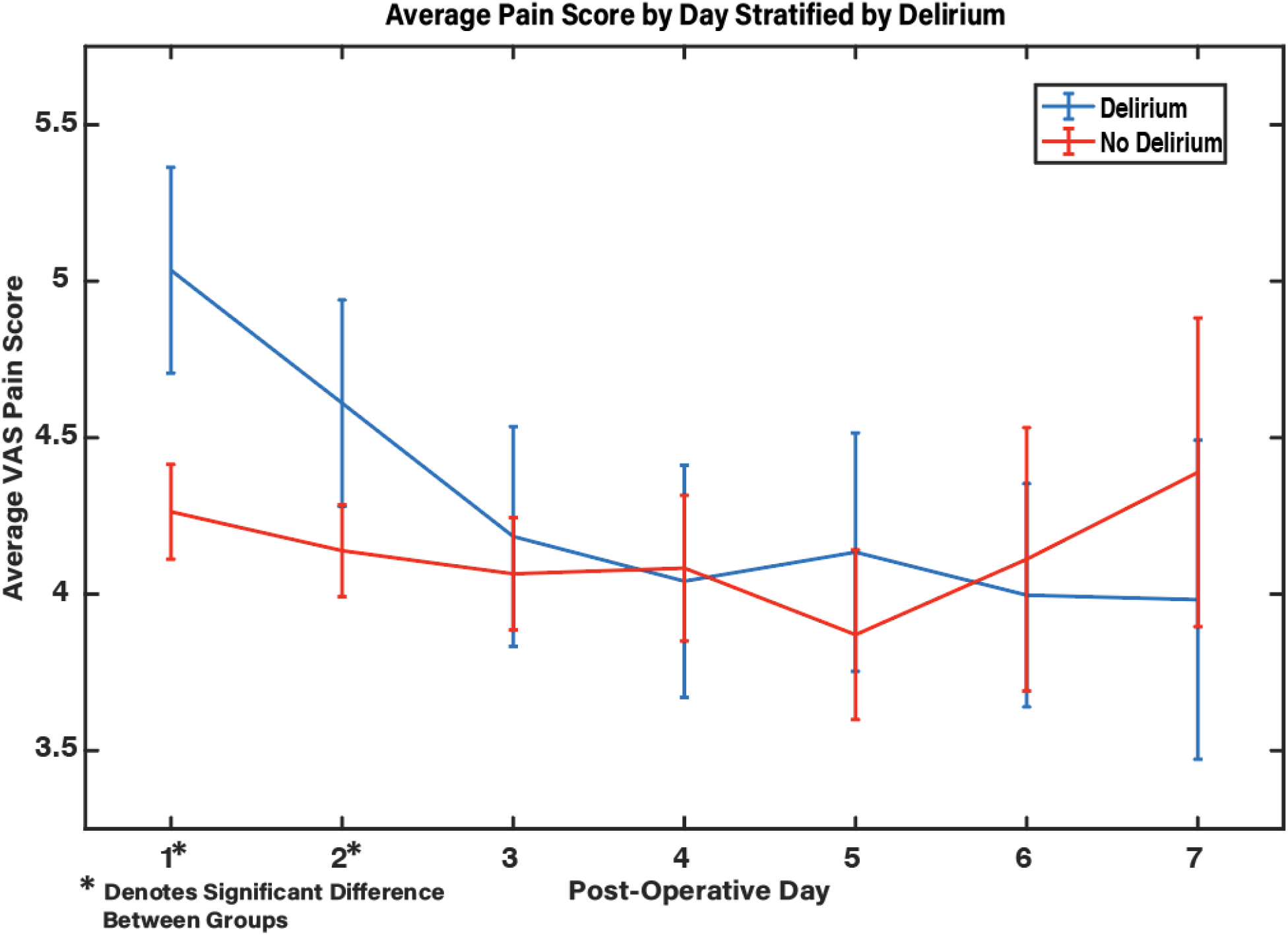
Line plot with 95% confidence intervals of average morphine equivalent dosage (MED) by postoperative day for delirious (blue) and non-delirious (red) patients. There was no significant difference between groups.

**Table 4.**
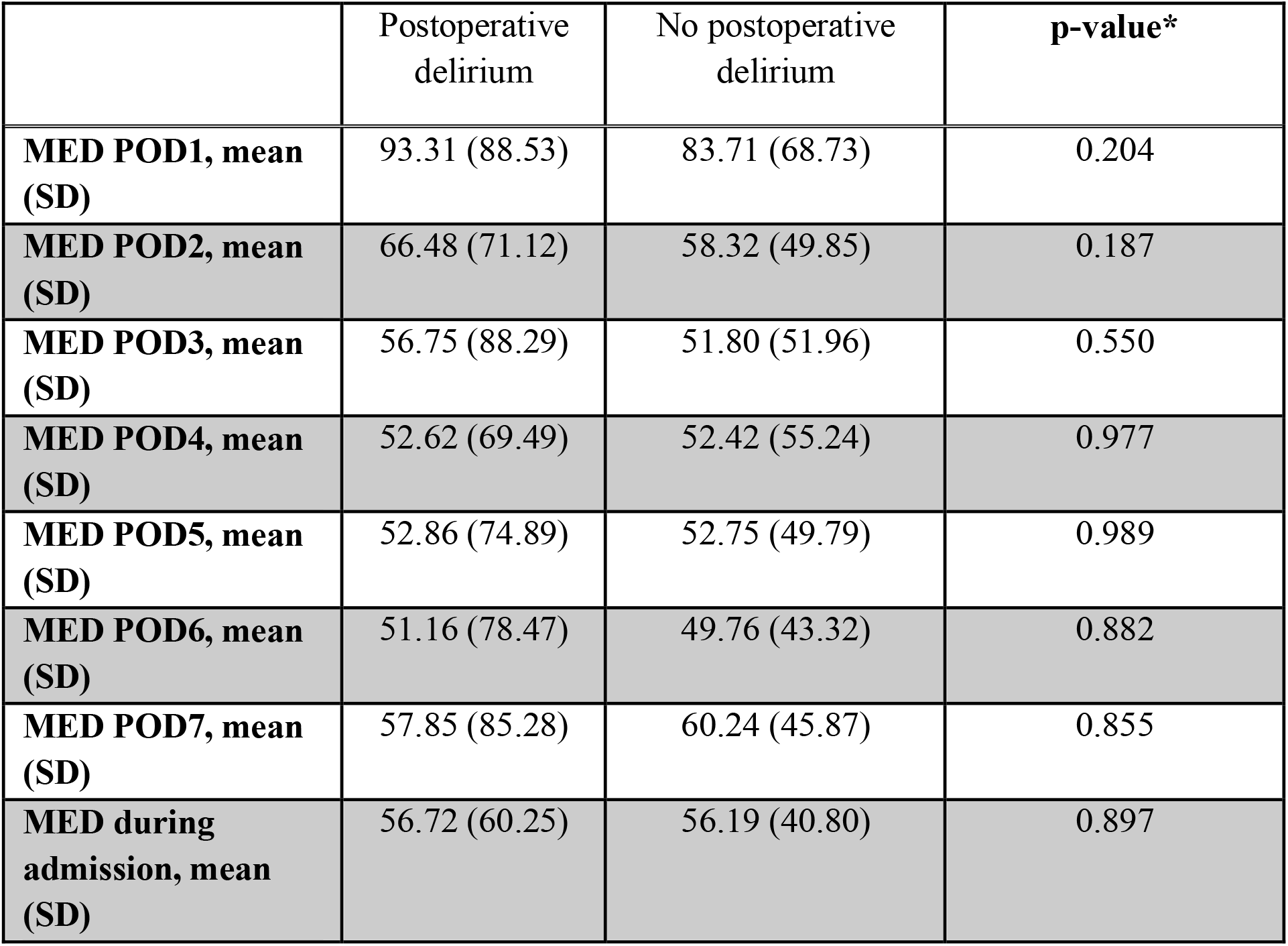
Mean Morphine Equivalent Dose (MED) compared between patients who developed delirium at any point during the postoperative period.

|  | Postoperative delirium | No postoperative delirium | **p-value\*** |
| --- | --- | --- | --- |
| **MED POD1, mean (SD)** | 93.31 (88.53) | 83.71 (68.73) | 0.204 |
| **MED POD2, mean (SD)** | 66.48 (71.12) | 58.32 (49.85) | 0.187 |
| **MED POD3, mean (SD)** | 56.75 (88.29) | 51.80 (51.96) | 0.550 |
| **MED POD4, mean (SD)** | 52.62 (69.49) | 52.42 (55.24) | 0.977 |
| **MED POD5, mean (SD)** | 52.86 (74.89) | 52.75 (49.79) | 0.989 |
| **MED POD6, mean (SD)** | 51.16 (78.47) | 49.76 (43.32) | 0.882 |
| **MED POD7, mean (SD)** | 57.85 (85.28) | 60.24 (45.87) | 0.855 |
| **MED during admission, mean (SD)** | 56.72 (60.25) | 56.19 (40.80) | 0.897 |

Informed by these separate univariate analyses, two separate pooled logistic regressions were performed to further assess the data and account for all covariates. Prior to this, all predictor variables were tested for collinearity – none of the pairwise comparisons reached a threshold for rejection of |r| ≥ 0.7.[19] In Model 1, independent variables included average VAS pain scores up to, and including, the day of onset of delirium, MED up to, and including, the day of delirium, age, gender, time under anesthesia, preoperative hemoglobin, diabetes, ASA status, EBL, and the number of spinal levels instrumented. Patients who became delirious were dropped from the model after the day they first became delirious. Overall, Model 1 significantly predicted postoperative delirium (*p* < 0.001) with an area under the curve (AUC) of 0.8079 (Figure 5; Table 5). Individually significant parameters within the regression Model 1 were postoperative day (OR 0.62, *p* < 0.001), average VAS (OR 1.32, *p* < 0.001), average MED (OR 0.84, *p* = 0.027), age (OR 1.07, *p* < 0.001), preoperative hemoglobin (OR 0.85, *p* < 0.001), and the ASA physical status (OR 1.54, *p* < 0.003).

**Figure 5.**
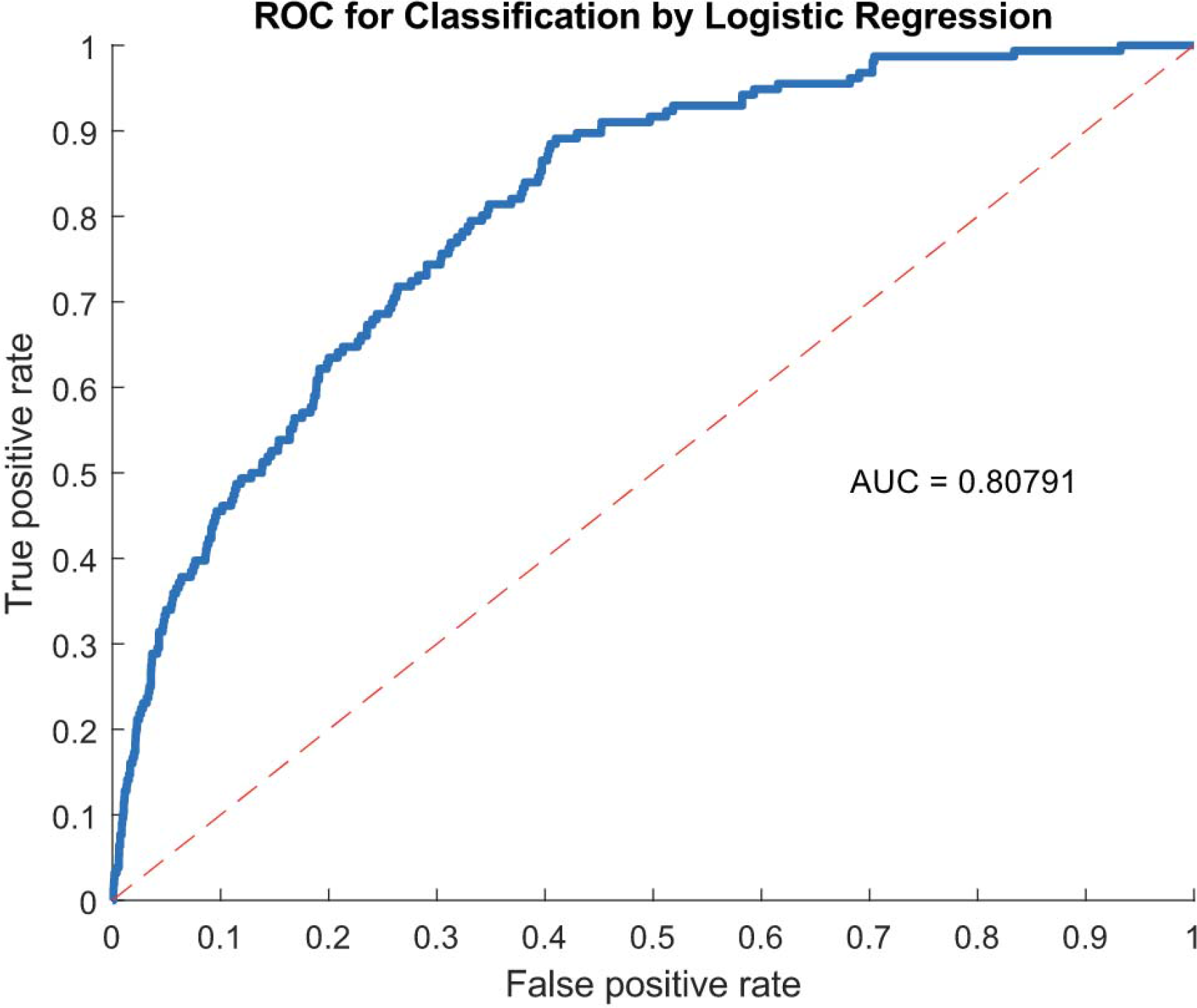
Receiver operating characteristic (ROC) curve for pooled logistic regression (Model 1) predicting postoperative delirium. The area under the curve (AUC) was 0.80791.

**Table 5:** Results of pooled linear regression model (Model 1) for predicting the presence of postoperative delirium, with data included up to and on the day of delirium of delirium onset.

|  | Estimate | SE | tStat | p value | Odds Ratio |
| --- | --- | --- | --- | --- | --- |
| **(Intercept)** | -10.05 | 2.86 | -3.51 | **<0.001** |  |
| **Day** | -0.48 | 0.06 | -8.62 | **<0.001** | 0.62 |
| **VAS Pain Score (mean)** | 0.28 | 0.06 | 4.93 | **<0.001** | 1.32 |
| **MED (mean)** | -0.18 | 0.08 | -2.21 | **0.027** | 0.84 |
| **Age** | 0.07 | 0.02 | 4.26 | **<0.001** | 1.07 |
| **Gender** | 0.16 | 0.18 | 0.86 | 0.388 | 1.17 |
| **Anesthesia time** | 0.33 | 0.29 | 1.15 | 0.249 | 1.39 |
| **Preoperative Hemoglobin** | -0.16 | 0.05 | -3.27 | **0.001** | 0.85 |
| **Diabetes** | 0.27 | 0.20 | 1.35 | 0.176 | 1.31 |
| **EBL** | 0.12 | 0.07 | 1.59 | 0.113 | 1.12 |
| **Levels Instrumented** | 0.04 | 0.03 | 1.22 | 0.223 | 1.04 |
| **ASA** | 0.43 | 0.17 | 2.48 | **0.013** | 1.54 |

To account for possible changes in pain scores and pain medications after the patient became delirious, a second model (Model 2) was created which included average VAS scores and MED prior to the onset of delirium (not including the day of onset of delirium); all other factors remained the same. Again, overall, Model 2 significantly predicted postoperative delirium (*p* < 0.001) with an area under the curve (AUC) of 0.7942 (Figure 6; Table 6). Similar to Model 1, the postoperative day (OR 0.63, *p* < 0.001), average VAS (OR 1.34, *p* < 0.001), preoperative hemoglobin (OR 0.87, *p* = 0.040), and age (OR 1.08, *p* < 0.001) were significant predictors for the presence of delirium. However, average MED (OR 1.04, *p* = 0.752) and ASA status (OR 1.48, *p* = 0.084) were no longer significant.

**Figure 6.**
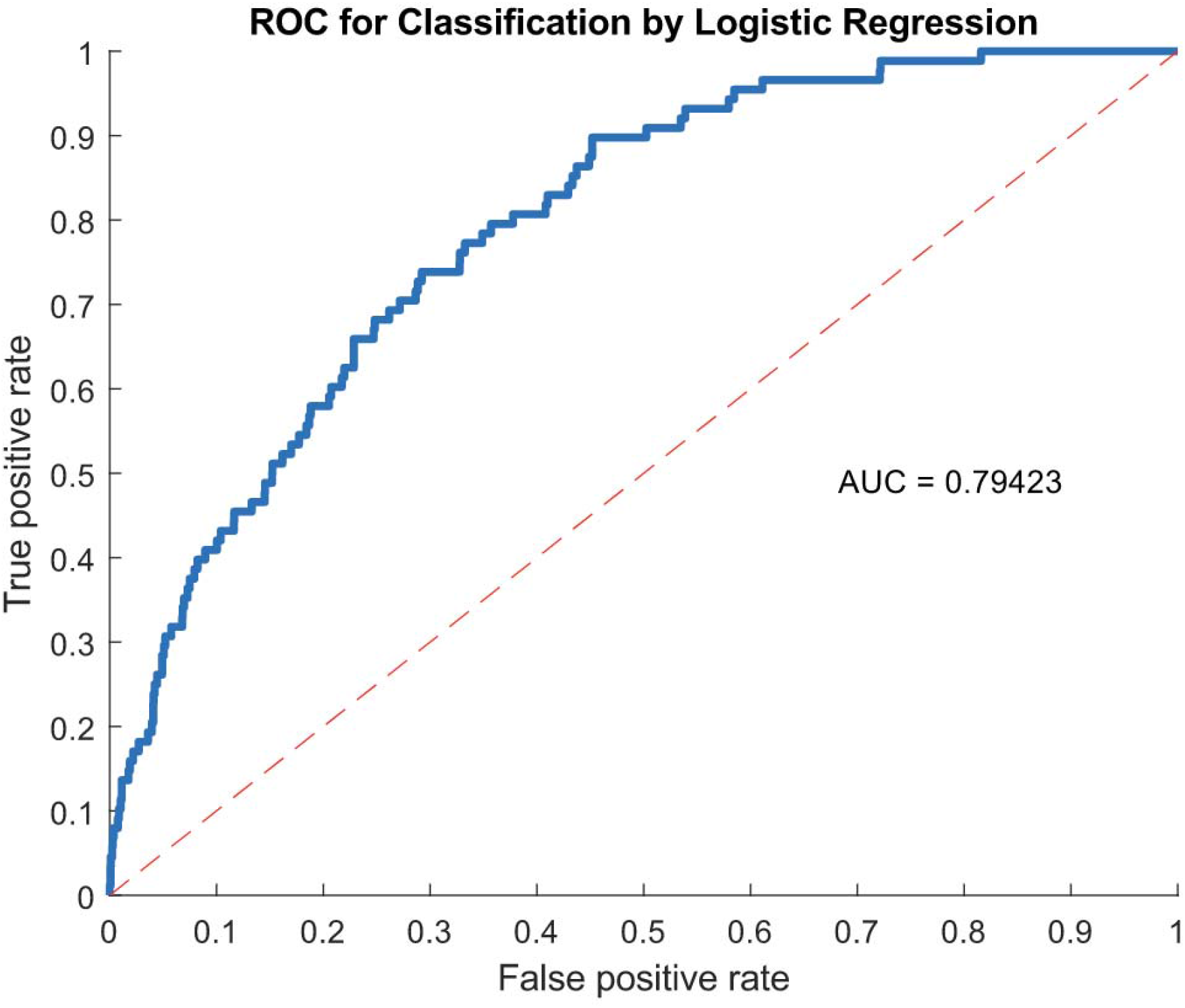
Receiver operating characteristic (ROC) curve for pooled logistic regression (Model 2) predicting postoperative delirium. The area under the curve (AUC) was 0.79423.

**Table 6:** Results of pooled linear regression (Model 2) for predicting the presence of postoperative delirium, with data included only when they were collected prior to the day of delirium onset.

|  | **Estimate** | **SE** | **tStat** | **p value** | **Odds Ratio** |
| --- | --- | --- | --- | --- | --- |
| **(Intercept)** | -12.91 | 3.73 | -3.46 | **<0.001** |  |
| **Day** | -0.46 | 0.08 | -5.73 | **<0.001** | 0.63 |
| **VAS Pain Score (mean)** | 0.29 | 0.07 | 3.89 | **<0.001** | 1.34 |
| **MED (mean)** | 0.06 | 0.12 | 0.48 | 0.633 | 1.06 |
| **Age** | 0.07 | 0.02 | 3.45 | **<0.001** | 1.08 |
| **Gender** | -0.15 | 0.24 | -0.61 | 0.545 | 0.86 |
| **Anesthesia time** | 0.48 | 0.36 | 1.32 | 0.186 | 1.61 |
| **Preoperative Hemoglobin** | -0.13 | 0.07 | -2.05 | **0.040** | 0.87 |
| **Diabetes** | 0.40 | 0.25 | 1.58 | 0.114 | 1.49 |
| **EBL** | 0.09 | 0.10 | 0.88 | 0.381 | 1.09 |
| **Levels Instrumented** | 0.06 | 0.04 | 1.59 | 0.113 | 1.06 |
| **ASA** | 0.27 | 0.23 | 1.20 | 0.230 | 1.32 |

## DISCUSSION

The incidence of delirium (24.6%) identified in this study is consistent with other reports of rates of delirium between 11-40% in patients after spine surgery[3, 4, 20-23]. The wide range of the reported incidence in the literature may be partly due to different inclusion criteria such as age, the type of spine surgery, use of instrumentation, the methods used for, and the frequency of, assessing for delirium. The average reported rate of postoperative delirium in the literature appears to be greater in prospective studies compared to retrospective ones (35% vs. 11%)[3, 4, 21-24] A meta-analysis including two prospective studies and three retrospective ones reported an incidence of 13%.[4] Moreover, delirium waxes and wanes^1^ and even twice daily assessments as performed at our institution could have missed delirious episodes. Therefore, given the fluctuating nature of delirium and the retrospective nature of this study, it is possible that the true incidence of postoperative delirium is higher than we observed. On the other hand, the screening instruments may overestimate the true prevalence of delirium. Validation of the DOSS in our hospital identified a positive predictive value (PPV) of 53% when compared to Delirium Rating Scale (DRS) scores ≥15. PPV increased to 94% when DRS scores suggesting subsyndromal delirium (8-14) was included.[18]

In the univariate analysis, significant preoperative risk factors for postoperative delirium were older age, higher ASA physical status, and a low preoperative hemoglobin. The presence of diabetes trended towards significance, however, with the correction for multiple comparisons it was not significant. The extent of surgery, as measured by number of levels of instrumentation, was significant, as was lower postoperative hemoglobin. More extensive surgery would cause more inflammation which has been proposed to be a triggering mechanism for delirium. High concentrations of peripheral inflammatory biomarkers have been shown to correlate with the presence of delirium.[4, 25] The majority (97%) of patients in this study were either ASA grade 2 or 3. The proportion of ASA 3 patients who developed delirium was significantly greater than that of ASA 2 patients (30.1% vs. 16.1%). Though not reaching statistical significance due to insufficient power, it is noteworthy that 40.0% of ASA grade 4 patients developed delirium. Our findings are consistent with other studies which have correlated the length of surgery, ASA status, and a higher intraoperative blood loss to the incidence of postoperative delirium.[26] Neither operative time nor anesthesia time as reported were found to be significant.

Other postoperative variables associated with post-operative delirium were admission to an ICU and the duration of stay in the ICU. Patients with POD had significantly longer times of hospitalization, were less likely to be discharged home, more likely to develop a surgical site infection, and were more likely to die within 90 days of surgery. Consistently, Ely *et al*. found that the presence of delirium increased the length of ICU stay and was the strongest predictor of hospital length of stay.[27] Patients with delirium have been reported to have a 62% increase in mortality during a one-year follow-up period.[9] Overall, our result are in agreement with those of other investigations of delirium in patients undergoing spine surgery. The cumulative data demonstrates worse clinical outcomes in patients with postoperative delirium,[3] creating a significant burden on patients, their families, healthcare, and society.

In the current study, average pain scores on the first day postoperatively were significantly associated with post-operative delirium. While overall average pain scores were not significant in a univariate analysis, both Model 1 and 2 in the pooled regression analysis found higher average pain scores to be predictive (OR 1.32, *p* < 0.001 and OR 1.34, *p* < 0.001 respectively). These findings are consistent with those in a study of older adults hospitalized for hip fractures, where undertreated pain and inadequate analgesia were found to be risk factures for delirium.[28] Another study that included all types of surgeries across several surgical specialties found that pain in the first three postoperative days was highly predictive of delirium, regardless of the method of analgesia, type of opioid used, and cumulative opioid dose.[29] However, to our knowledge, this is the only study in strictly lumbar spinal fusion patients linking higher postoperative pain to delirium.

To investigate whether pain itself was causing delirium or whether the medication used to treat pain triggered delirium, we reviewed analgesic medications including the daily use of opioids (MED) and perioperative use of gabapentin. There was no difference in the incidence of delirium in patients who received gabapentin at any time during their hospital stay. However, patients who suffered from delirium received a significantly higher total dosage of gabapentin over the course of the hospital stay. In the univariate analysis, there were no significant differences between MED in delirious and non-delirious patients. However, in the pooled regression, lower average MED was a significant predictor of delirium when the day of delirium onset was included (Model 1). But this was not the case when the day of delirium onset was excluded (Model 2). One likely explanation for these differences is that usually, after delirium is diagnosed, medications that could influence the cognitive status are withheld. Furthermore, patients with altered mental status may not be able to express that they have pain and ask for analgesic medication. Since twice daily assessments had been conducted, we estimated the time scale of delirium using the postoperative day rather than postoperative hours. Hence it is possible that the onset of delirium preceded cessation of narcotic medications resulting in a significantly lower average MED; however, this cannot be established by our data.

This study adds to a body of evidence suggesting that early, uncontrolled postoperative pain increases the risk of delirium. To date, there is a lack of consensus about the optimal method of pain control in these patients. For example, some have suggested that oral opioids are superior to opioids when delivered by IV patient-controlled analgesia.[30] Rodents that underwent surgery and received inadequate postoperative analgesia developed greater acute cognitive dysfunction and showed evidence of an increased amount of inflammatory cytokines in the prefrontal cortex and hippocampus at autopsy.[31]

To reduce the incidence of postoperative delirium, a fundamental necessity is to understand the factors that cause and/or exacerbate it so that mitigating strategies can be incorporated into perioperative care pathways. The current findings warrant future investigation into the effect of preventing and/or treating perioperative anemia as well as that of improved postoperative analgesia on postoperative delirium in elderly patients after posterior lumbar spinal fusion. Additionally, investigation of the link between pain and delirium using neurophysiological data e.g. inflammatory markers in the cerebrospinal fluid, neuroimaging, and electroencephalography, could lead to significant discoveries about the currently poorly understood phenomenon of postoperative delirium.

### Limitations

We must acknowledge the following limitations in this study. First, this was a retrospective cohort study. Differences in surgical techniques and perioperative management between the Neurosurgery and Orthopedic surgeons and other staff in our hospital may have confounded the results. Patient reported outcomes (PROs) such as Oswestry Disability Index (ODI) or Patient-Reported Outcomes Measurement Information System (PROMIS) are one way to standardize metrics between departments and surgeons. Unfortunately, these PROs were not uniformly collected at our institution during the period of review.

Delirium scores were determined using the DOSS which has been shown to be less specific than other measures such as the CAM-3D.[32, 33] Additionally, the reasons for admission to the ICU were not collected. It is possible that in some cases, delirium could have been triggered by postoperative events such as sepsis. We did not collect data on baseline pain scores, all preexisting co-morbidities and the use of other medications, both chronically consumed and received in the perioperative period including anesthetic drugs. Preoperative opioid and anti-depressant use, and preoperative cognitive dysfunction (MMSE) use have also been associated with increased rates of delirium.[3, 4, 22, 26, 34] However, this data was either not available (drugs) or there were not a sufficient number of patients in the study cohort (preoperative cognitive dysfunction).

## CONCLUSION

A retrospective review of data from elderly patients undergoing posterior lumbar spinal fusion at our institution characterized the incidence of, risk factors for, and outcomes due to, postoperative delirium. The incidence of post-operative delirium in 702 patients undergoing elective posterior lumbar fusions was found to be 24.6%. Univariate analysis showed that age, low preoperative hemoglobin, and higher ASA scores were significantly associated with POD. Intraoperatively, the number of instrumented spinal levels, and a lower postoperative hemoglobin correlated with a higher incidence of POD. Postoperative admission to the ICU and increased length of ICU stay, a higher total gabapentin dosage, and greater pain in the first postoperative day were associated with an increased incidence of delirium. Patients who experienced delirium had significantly longer lengths of hospitalization, were less likely to be discharged home, were more likely to develop a surgical site infection, and had higher rates of mortality within 90 days after surgery. Regression models identified average pain scores up to the time of onset of delirium, age, and the ASA status as potential key factors predictive of POD. This study provides further understanding of the complex etiology of POD and provides an institutional benchmark for future studies into the mechanisms of POD, and interventions to prevent and/or reduce its severity.

## Data Availability

All data produced in the present study are available upon reasonable request to the authors except data that is HIPAA protected.

## Acknowledgements

Thank you to Scott Seaman MD for his assistance with the statistical analysis. Thank you to Gregory Hopson and Linda Kleinmeyer for their assistance with the acquisition of the data from the electronic health records.

## Abbreviations

(POD): Postoperative Delirium

